# Assessing the completeness of reporting of exercise-based interventions in cardiovascular trials

**DOI:** 10.1101/2022.06.06.22276052

**Authors:** GW Freccia, RZ Santos, L Lucca, AS Korbes, T Carvalho

## Abstract

**Study Objective:** In cardiology, the reporting of interventions is insufficiently detailed to elicit replication. Specifically for cardiovascular rehabilitation (CVR) trials, the quality of the description of its exercise-based interventions (EBI) is poorly known. Our primary objective is to estimate the adherence of EBI of CVR trials to TIDieR reporting guideline. Secondarily, we tested whether the CVR setting would be associated with a better reporting, also exploring whether transparency and methodological characteristics could be related.

**Design and Setting:** This cross-sectional study analysed 96 trials with EBI published within CVR literature. For our primary objective, TIDieR reporting guideline was accounted as a reference to assess overall adherence of eligible RCTs. For our secondary objectives we used generalised estimating equations to point out (a) if intervention setting (eg, home-based vs. centre-based) was associated with intervention reporting, and (b) if trials transparency and methodological characteristics would be associated with intervention reporting.

**Results:** On average, arms adequately reported 4.8/12 (SD=2.4) TIDieR items. 65.07% of our EBI arms failed to adequately report ≥ 6 TIDieR items. Three of 146 (2.05%) arms adhered to all 12 TIDieR items. Additionally, intervention setting was not associated with a better description.

**Conclusions:** We concluded that EBIs in CVR lack to report fundamental aspects of the interventions to be replicated by third parties based on their reports.

**Highlights**

1. We found an inadequate and insufficient exercise-based interventions reporting of cardiovascular rehabilitation trials.
2. Overall, about 40% of TIDieR reporting guideline adherence was found, with most interventions reporting its items improperly.
3. Exercise-based intervention care setting (home- and centre-based) have no association with a better intervention description.
4. Studies with 2 or more EBI arms are associated with a poor TIDieR adherence.
5. Publishers and Editorial committees may guide cardiology journals for a better intervention reporting.

## 1. Introduction

Cardiovascular rehabilitation programs (CVR) are crucial for the management of cardiovascular diseases (CVD), impactfully reducing its morbimortality [1]. The evidence-based medicine is also a cornerstone for an adequate patient care [2]. To reach this goal, one of the componentes of the knowledge translation construct is the adequate reporting of interventions [3].

Meta-research studies problems and solutions to improve research [4], providing empirical evidence regarding problems like the impact of insufficient or overestimated samples [5], the problems of small effect sizes [6], poorly reproducible methods and results [7], and poor adherence to guidelines [3,8]. Ultimately, a basic assumption of science needs to be warranted: the capacity of a scientific report to be reproducible by methods, results, or inferences.

The level of reproducibility in Cardiology is still unknown [9], with very few evidence suggesting inconsistencies and methodological weaknesses within its randomized clinical trials (RCTs) [7,10–12]. A reproducible study must be sufficiently detailed and accurate to elicit readers and independent researchers to replicate. For parallel-RCTs, the CONSORT 2010 reporting guideline helps researchers to report their experiments in an adequate manner [13]. In addition, an extension of this guideline focusing only on the interventions section is available - the TIDieR reporting guideline [14]. Among the exercise-based intervention (EBI) CVR studies, about half provide an adequate reporting of interventions [3], without relevant improvement over time regardless of the scientific community effort and editorial policies to improve the quality of reporting of manuscripts [15].

Given this context, we aimed to estimate the adherence of CVR interventions based on physical exercise to TIDieR guideline, as our primary objective. Secondarily, we tested whether the mode of CVR intervention’s care setting (*e.g*., home-based, centre-based, etc.) would be associated with adherence to the TIDieR. We also explored whether potential variables from transparency and methodological characteristics domains could be independently related with the level of adherence to TIDieR reporting guideline.

## 2. Methods

This is a cross-sectional survey of the literature of CVR RCT. We reported this study based on the recommendations of the reporting guideline proposed by Murad and Wang [16]. The protocol, materials, statistical analysis scripts and raw data are publicly accessible in our OSF repository (<u>link</u>).

### 2.1. Eligibility criteria

RCTs met the following criteria to be eligible: (a) a CVR trials containing at least one arm including any type of EBI; (b) and randomized patients with CVD-related morbidities, as follows: coronary artery disease, stable angina, unstable angina, acute myocardial infarction (STEMI and non-STEMI), heart failure independente of the ejection fraction status, primary/secondary hypertension, cerebrovascular disease (ischemic or hemorrhagic stroke), peripheral arterial disease.

EBI CVR was defined as a synonymous of “cardiac rehabilitation” that should be composed by physical exercise components, whether being or not a multicomponent intervention [17]. In our study, EBI consisted at least by aerobic or resistance exercises, supervised or not, individually or in groups, delivered at home (home-based) or another setting, in patients with diagnosed CVD [18]. We considered inspiratory muscle training and whole-body electrostimulation training as eligible interventions, as both of them can elicit strenght against an external resistance and therefore result in chronical adaptations in physical fitness [19,20]. Studies with arms in which primary intervention was motivational, nutritional, counseling, or usual care (*i.e*., standard medical care such as medication use or a non-pharmacological, non-structured intervention) were not eligible. Usual care arms were not included in our analysis, as they may differ greatly regarding their definitions and contexts.

### 2.2. Search strategy and screening

To gather our unit of analysis systematically there was a sequential step process, starting by surveying journals of interest, and then applying the search strategy in each of them, to therefore retrieve all eligible manuscripts for analysis. We first queried Scopus database to gather journals of interest using the “*Journals*” filter, namely as strata: “*Physical Therapy, Sports Therapy and Rehabilitation*”, “*Cardiology and Cardiovascular Medicine*”, and “*Rehabilitation*”.

After that, we took the top 100 journals ranked by Scopus CiteScore 2019, which were screened by a single author (GWF) to indicate those that met and exclude those that did not meet our eligibility criteria: (a) scope of interest (*i.e*., Cardiology and Rehabilitation) and (b) have at least one CVR parallel-RCT published between 2017 onwards. The journals’ scope (*e.g*., Orthopedic Rehabilitation, Surgery, Imaging) or design (*e.g*., Review Studies, Basic or Experimental Research, *etc*.) that did not meet our eligibility criteria were deemed as ineligible, as well as those not published in English, Spanish, Portuguese, and French, for reasons of feasibility.

We then searched for RCTs of CVR in each eligible journal applying a search strategy in PubMed/MEDLINE, composed by free terms, MeSH terms, and relevant descriptors related to exercise, plus an ultrasensitive filter for RCTs (Appendix A) [21]. 10% of the retrieved articles were piloted to test the eligibility process flow, as well as library management. After the approval of a senior researcher about the pilot sampling, the retrieved articles were organized in Zotero 5.0.96 reference manager software by GWF.

### 2.3. Study selection

Retrieved articles were organized alphabetically by title and were deduplicated with Zotero plus a manual checking. The study selection process was conducted in a duplicate of independent and blinded reviewers (GWF + LL and ASK + RZ), from the titles and abstracts. Disagreements were adjudicated by a third investigator (LH). Reasons for exclusions were registered in a Google Sheets document (accessible on our repository). The flowchart with the eligibility process of the journals and articles selected for the sample is shown in **Figure A**.

**Figure A.**
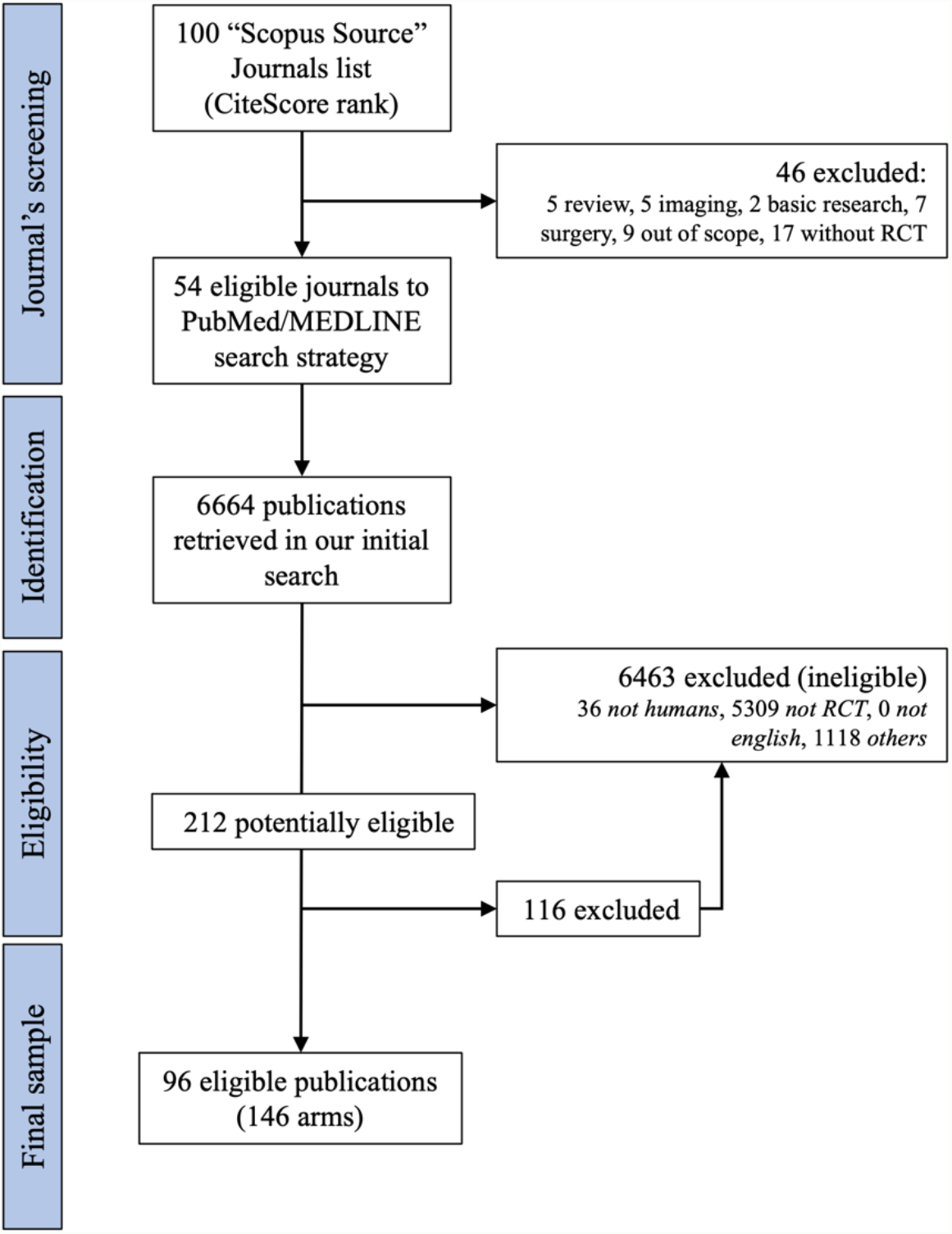
Eligibility process flowchart of the journals and articles. RCT: Randomized Controlled Trial; EBI: Exercise-based intervention

### 2.4. Data extraction and training

Prior to data collection, a form was created in Google Forms, piloted and, after approval of an experiencied researcher (TC), we moved forward to data extraction. A single author (GWF) extracted an article [22] to conduct the training sessions with the data extraction team through Microsoft Teams to establish consistency within the extraction. The training consisted of a review and explanation of the project’s objectives, methods, and outcomes, as well as the structure of the previously designed form. The record of the training session (audio in PT-BR) as well as the form and extraction worksheets are available in our public repository.

The data extraction was carried forward in the same manner and by the same researchers after the screening phase. Disagreements were solved by consensus. We extracted the following items: (a) number of EBI arms; (b) year of publication; (c) impact factor (SJR); (d) corresponding author’s country; (e) journal’s continent; (f) material’s sharing; (g) data availability; (h) analysis scripts availability; (i) protocol; (j) study (pre)registration^1^; (k) whether it’s a replication or a novel study; (l) conflict of interest declarations; (m) funding sources disclosure; (n) open access article accessible link. These variables were deemed as relevant to be gathered based on previous studies, as they may be associated to the adherence to reporting guidelines [23,24].

TIDieR reporting guideline was taken as the main reference to the completeness of reporting of eligible RCTs. Whenever a TIDieR item was properly reported in the primary study, we assigned as YES (value = 1) to attend the purporse of this study, otherwise we assigned as NO (value = 0). Additional instructions were provided additionally to the main TIDieR items (3 to 12), adapting the TIDieR reporting guideline to cope with the specificities of EBIs [25], which can be found on Appendix B. Missing or insufficiently described intervention items were considered incomplete.

To answer our secondary outcomes, the rehabilitation setting was also extracted, categorized as: home-based rehabilitation, centre-based (*e.g*., clinic, hospital, laboratory) rehabilitation, or the combination of both (independent of the order). Both studies and arms sample size were retrieved, as well as the targeted sample size (ie, sample size calculation). These additional variables were collected by a single investigator (GWF) after our first protocol version.

### 2.5. Statistical analysis

Descriptive statistics for the primary outcome were described as counts and prevalences for categorical variables, within a 95% of precision of estimates (95% confidence intervals – 95%CI). For continuous variables, we described estimates through means and standard deviations (SD) or medians and interquartiles intervals (range or minimum-maximum), dependending of its density distribution.

Our primary goal was to assess the completeness of reporting of EBI in the RCTs as measured by adherence to TIDieR items (total point-completion as YES). It is expected that the editorial policies of a very same jornal have a similar impact in all the publication output of the journal, although we may consider the role of referees, handling editors or even personal decisions that may distort a pattern to be expected. Despite of what was ackonowledged in the end, it is fair to expect a Poisson distribution for the completeness of reporting, which was found when exploring our histogram. Then, we regressed the adherence to TIDieR items with the intervention delivery mode through a crude Poisson regression model. To deal with transparency- and methodological-related predictors, we regressed the same outcome by a multivariate negative binomial regression model accomodated in a generalized estimating equation model (GEE), with a time-variable equal to one endpoint only (a way to command the longitudinal analysis as a cross-sectional analysis). The random effect function was inserted in the model with the identity link function, as well as an independent correlation structure [26].

Prevalence ratios (PR) were generated and approximated for relative risks in outputs for interpretation [27,28]. In addition, robust variances were considered for the model in Poisson regression. All analysis were conducted in Stata 16.0.0 and R 4.0.1 GUI 1.72, and scripts (command lines) are available in our public repository.

## 3. Results

### 3.1. Demographic, general, and methodological characteristics

From our sample of studies (N=96), 54.17% (52/96; 95%CI 44.04-63.96) of RCTs have only one EBI study group arm. 38.54% (37/96; 95%CI 29.26-48.74) have two and 7.29% (7/96; 95%CI 3.48-14.64) has three EBI arms. 20.83% (of the studies come from United States of America (USA), followed by 10.42 % from Brazil, 7.29% from Australia, 7.29% from the Netherlands, 5.21% from China, and 5.21% from France, distributed between Europe (55.21%) and Americas (42.71%).

The mean SJR plus standard deviation was 2.35±1.89 (95%CI 1.95-2.56). In terms of randomized sample size, RCTs ranged from N=12 up to N=1984, within a median of N=73. 43.75% (95%CI 34.08-53.92) randomized less then 50 patients. 87.50% (84/96; 95%CI 79.14-92.81) of the studies declared in the article whether there was a source of funding or not. 78.12% (75/96; 68.63-85.36) have some mention about potential conflicts of interest; and 61.46% (59/96; 51.26-70.74) were published in open access mode. On the same manner, 76.04% (73/96; 95%CI 66.38-83.61) of the RCTs reported to have been prospectively registered; and 16.67% (16/96; 95%CI 10.40-25.62) provide a study protocol. Whenever evaluated about their characteristics of practices for research transparency and individual participant data (IPD) sharing, 36.46% (35/96; 95%CI 27.36-46.64) of the studies disclosed any type of data availability, and 3.12% (3/96; 95%CI 1.00-9.37) shared the raw data (or reported that they would be available under request). Remaining characteristics can be seen at **Table 1**.

**Table 1.** General and demographic characteristics from our studied sample.

| Characteristics (N=96) | Reference | N (%) | 95%CI |
| --- | --- | --- | --- |
| How many EBI arms does the study have? | 1 | 52 (54) | 44.0 - 63.9 |
|  | 2 | 37 (38) | 29.3 - 48.7 |
|  | 3 | 7 (7) | 3.5 - 14.6 |
| What was the publication date? | 2017 | 26 (27) | 19.1 - 36.9 |
|  | 2018 | 18 (18) | 12.1 - 27.9 |
|  | 2019 | 22 (22) | 15.5 - 32.5 |
|  | 2020 | 27 (28) | 19.9 - 38.0 |
|  | 2021 | 3 (3) | 1.0 - 9.3 |
| Journal Impact Factor (SJR) | Mean | 2.35 | 1.9 - 2.5 |
|  | SD | 1.89 |  |
|  | Min-Max | 0.79 - 10.31 |  |
| What is the country of the corresponding author? | Australia | 7 (7) |  |
|  | Germany | 5 (5) |  |
|  | Brazil | 10 (11) |  |
|  | China | 5 (5) |  |
|  | USA | 20 (21) |  |
|  | France | 5 (5) |  |
|  | Netherlands | 7 (7) |  |
|  | Others | 37 (39) |  |
| What is the journal's continent? | Americas | 41 (42) | 45.1 - 64.9 |
|  | Europe | 53 (55) | 33.1 - 52.9 |
|  | Oceania | 2 (2) | 0.5 - 8.1 |
| RCT sample size | Mean | 145.6 | 92.7 - 198.5 |
|  | Median | 73 |  |
|  | Min - Max | 12 - 1984 |  |
|  | p5% - p95% | 13 - 914 |  |
| RCT sample size (categorized) | < 50 | 42 (44) | 34.1 - 53.9 |
|  | 51 - 100 | 19 (20) | 12.9 - 29.1 |
|  | 101 - 200 | 20 (21) | 13.8 - 30.2 |
|  | 201 - 300 | 7 (7) | 3.4 - 14.6 |
|  | > 301 | 8 (8) | 4.1 - 15.9 |
| Arm size | Mean | 61.82 | 46.6 - 77.0 |
|  | Median | 28 |  |
|  | Min - Max | 6 - 580 |  |
|  | 5% - 95% | 6 - 425 |  |
| Does the article state whether materials are available? | No | 71 (74) | 64.1 - 81.8 |
|  | Yes | 25 (26) | 18.1 - 35.8 |
| Does the article state whether data are available? | No | 61 (64) | 53.3 - 72.6 |
|  | Yes | 35 (36) | 27.3 - 46.6 |
| Does the article state whether analysis scripts are available? | No | 93 (97) | 90.6 - 99.0 |
|  | Yes | 3 (3) | 1.0 - 9.3 |
| Does the article have a link to an accessible protocol? | No | 80 (83) | 74.3 - 89.6 |
|  | Yes | 16 (17) | 10.4 - 25.6 |
| Does the article state whether the study (or some aspect of the study) was pre-registered? | No | 23 (24) | 16.4 - 33.6 |
|  | Yes | 73 (76) | 66.4 - 83.6 |
| Does the article claim to report a replication study or novel study? | No | 76 (79) | 69.8 - 86.2 |
|  | Yes, novel | 20 (21) | 13.8 - 30.2 |
|  | Yes, replication | 0 (0.00) |  |
| Does the article include a statement indicating whether there were any conflicts of interest? | No | 21 (22) | 14.6 - 31.4 |
|  | Yes | 75 (78) | 68.6 - 85.4 |
| Does the article include a statement indicating whether there were funding sources? | No | 12 (13) | 7.2 - 20.8 |
|  | Yes | 84 (87) | 79.1 - 92.9 |
| Is the article open access? | No | 37 (39) | 29.3 - 48.7 |
|  | Yes | 59 (61) | 51.3 - 70.7 |

**Note:** EBI: exercise-based intervention; RCT: randomized clinical trial; SJR: Scimago Journal Rankings; SD: standard deviation.

### 3.2. Primary outcome

Of 12 items, on average, arms adequately reported 4.8±2.4 items from TIDieR. 65.07% of our EBI arms failed to report ≥ 6 TIDieR items. Three of 146 (2.05%) arms adhered to all 12 TIDieR items. **Figure B** presents a histogram that illustrate the frequency of adherence of TIDieR items (**note**: arms as unit of analysis).

**Figure B.**
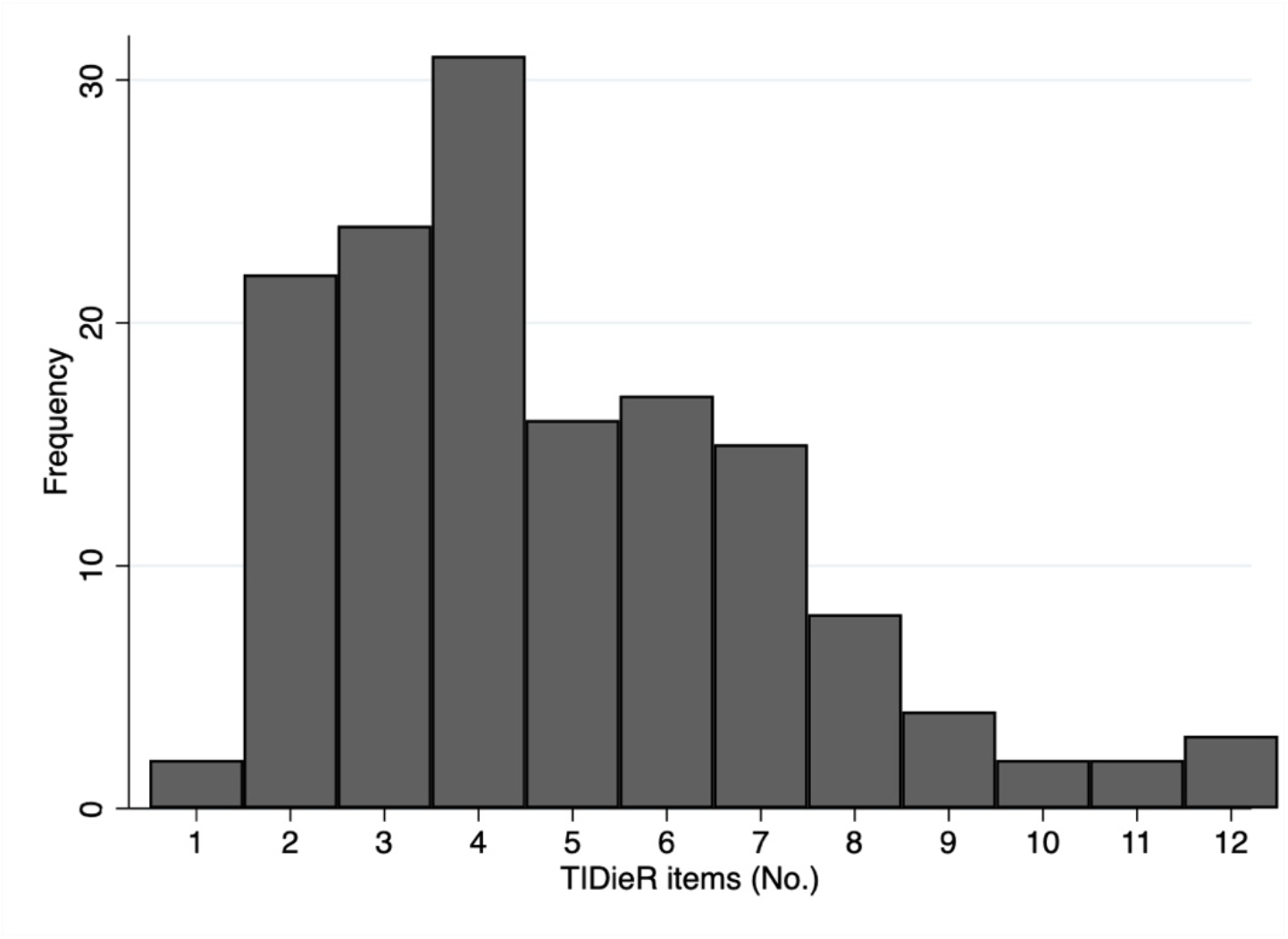
Distribution of arms by the number of reported TIDieR items.

Of the items, “*Brief Name*” and “*Why*” had 97.2% and 95.21% of adherence respectively. Items likely “*Where*” (16.44%), “*Tailoring*” (25.34%), “*Modifications*” (6.16%), “*How well: planned*” (20.55%), and “*How well: actual*” had an adherence of 16.44%, 25.34%, 6.16%, 20.55% and 16.44% respectively. EBI arms adherence item-by-item are fully described in **Table 2**.

**Table 2.** TIDieR item-by-item adherence given the analyzed arms.

| TIDieR Item<br>N=146 | Variables<br>% (95%CI) |  |
| --- | --- | --- |
| **(1) Brief Name**<br>Do the authors provide the name or a phrase that describes the intervention? | No<br>Yes | 2.7 (0.7 - 6.8)<br>97.3 (93.1 - 99.2) |
| **(2) Why**<br>Do the authors describe any rationale, theory, or goal of the elements essential to the intervention? | No<br>Yes | 4.8 (1.9 - 9.6)<br>95.2 (90.3 - 98.0) |
| **(3) What: materials**<br>Do the authors describe any physical or informational materials used in the intervention, including those provided to participants or used in intervention delivery or in training of providers, or provide information on where the materials can be accessed (online appendix or URL)? | No<br>Yes | 53.4 (45.0 - 61.7)<br>46.6 (38.3 - 55.0) |
| **(4) What: procedures**<br>Do the authors describe each of the procedures, activities and/or processes used in the intervention, including any enabling or support activities? | No<br>Yes | 56.8 (48.4 - 65.0)<br>43.2 (35.0 - 51.6) |
| **(5) Who Provided**<br>For each category of intervention provider (psychologist and nursing assistant), describe their expertise, background and any specific training given? | No<br>Yes | 69.9 (61.7 - 77.2)<br>30.1 (22.8 - 38.2) |
| **(6) How**<br>Do the authors describe the modes of delivery (face-to-face or by some mechanism, such as internet or telephone) of the intervention and whether it was provided individually or in a group? | No<br>Yes | 61.0 (52.5 - 68.9)<br>39.0 (31.0 - 47.4) |
| **(7) Where**<br>Do the authors describe the type(s) of location(s) where the intervention occurred, including any necessary infrastructure or relevant features? | No<br>Yes | 83.6 (76.5 - 89.1)<br>16.4 (10.8 - 23.4) |
| **(8) When and How Much**<br>Do the authors describe the number of times the intervention was delivered and over what period including the number of sessions, their schedule, and their duration, intensity or dose? | No<br>Yes | 52.1 (43.6 - 60.4)<br>47.9 (39.6 - 56.3) |
| **(9) Tailoring**<br>If the intervention was planned to be personalised, titrated, or adapted, do the authors mention so and then describe what, why, when and how? | No<br>Yes | 74.7 (66.8 - 81.4)<br>25.3 (18.5 - 33.2) |
| **(10) Modifications** | No | 93.8 (88.6 - 97.1) |
| Do the authors mention if the intervention was modified during the study, and if so, describe the changes (what, why, when, and how)? | Yes | 6.2 (2.8 - 11.3) |
| **(11) How well: planned** | No | 79.4 (72.0 - 85.7) |
| If intervention adherence of fidelity was assessed, do they describe how and by whom, and if any strategies were used to maintain or improve fidelity, did the authors describe them? | Yes | 20.6 (14.3 - 28.0) |
| **(12) How well: actual** | No | 83.6 (76.5 - 89.1) |
| (If the above answer was yes) If intervention adherence or fidelity was assessed, did the authors describe the extent to which the intervention was delivered as planned? | Yes | 16.4 (10.8 - 23.4) |

### 3.3. Secondary outcomes

To estimate the association between TIDieR items adherence and intervention setting, we used a Poisson regression model with robust variances accommodated in a general linear model, without adjustments. We found the following prevalence ratios (PR) for centre-based delivered and home-based delivered interventions – PR=1.06 (95%CI 0.74-1.52, N=146; *P*=0.73) and a PR=1.13 (95%CI 0.76-1.66; N=146; *P*=0.53), considering the centre+home-based as the category of reference.

We also explored if variables related to transparency practices and methodological aspects of RCTs would be associated with TIDieR adherence of RCTs. For the number of study arms, those with three EBI arms had a PR=0.74 (95%CI 0.55-0.92; *P*=0.010) and the ones of two a PR=0.84 (95%C 0.71-0.99; *P=*0.041), against only one arm of EBI.

Whenever considering continents as potential predictors of TIDieR items adherence, RCTs published in journals from Americas had a PR=1.29 (95%CI 1.06-1.57; *P*=0.010) and those from Oceania a PR=1.99 (95%CI 1.33-2.97; *P*=0.001), against Europe. Among transparency characteristics, materials availability had an independent with TIDieR adherence, mirrored by a PR=1.31 (95%CI 1.11-1.54; *P*=0.001). Full crude and adjusted analysis are displayed in **Table 3**.

**Table 3.**
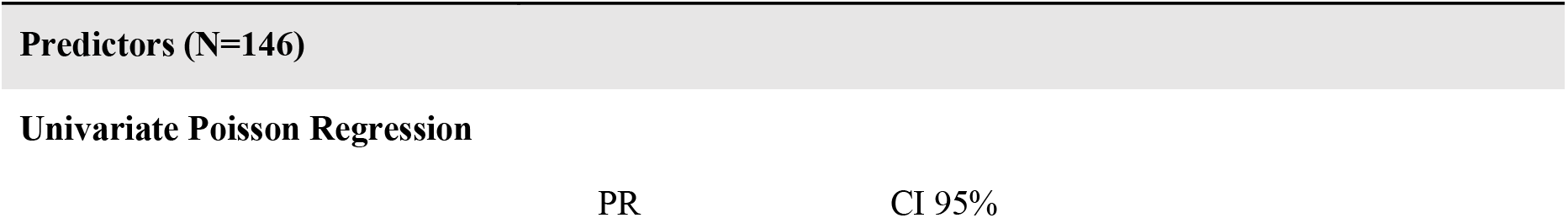
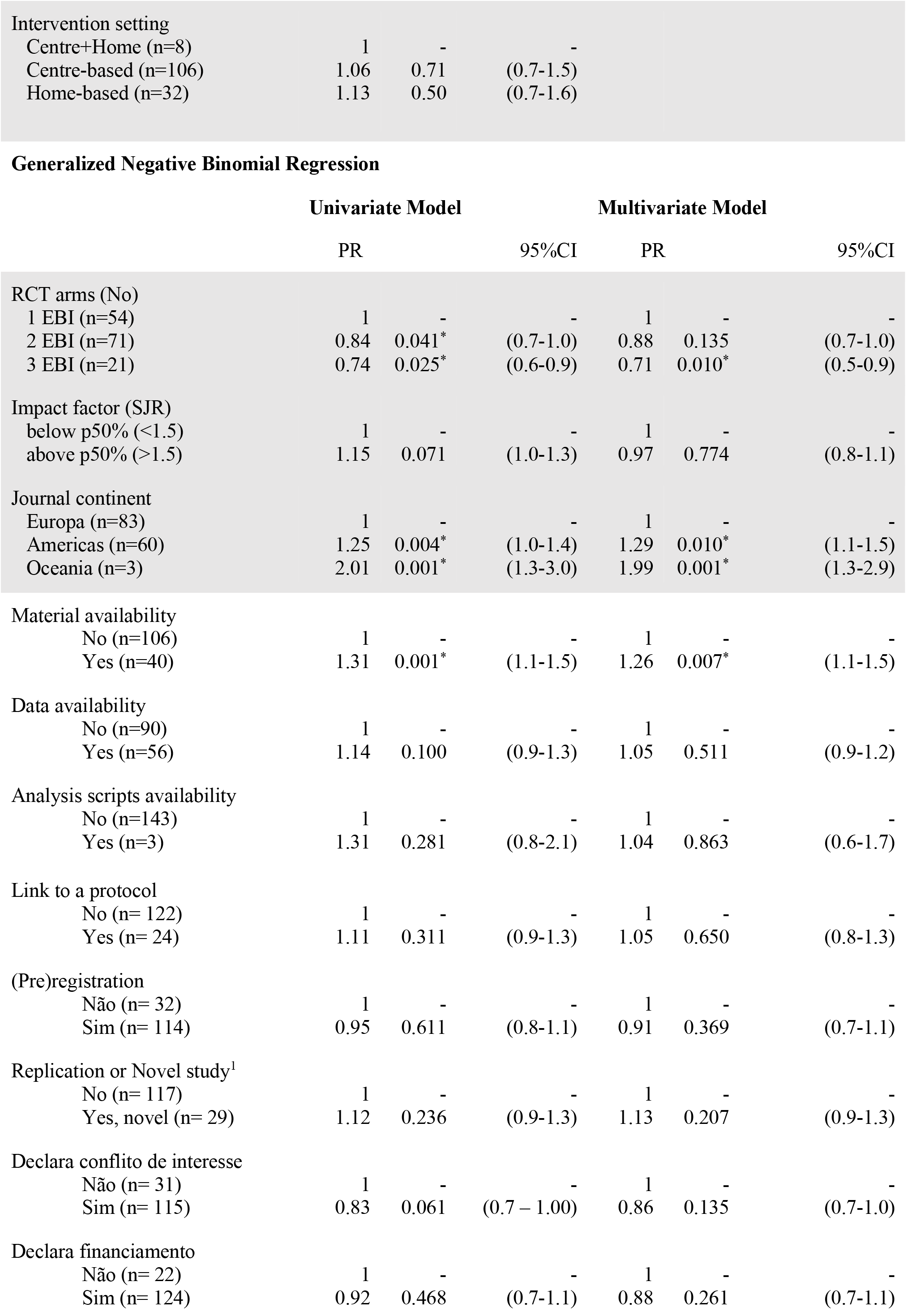
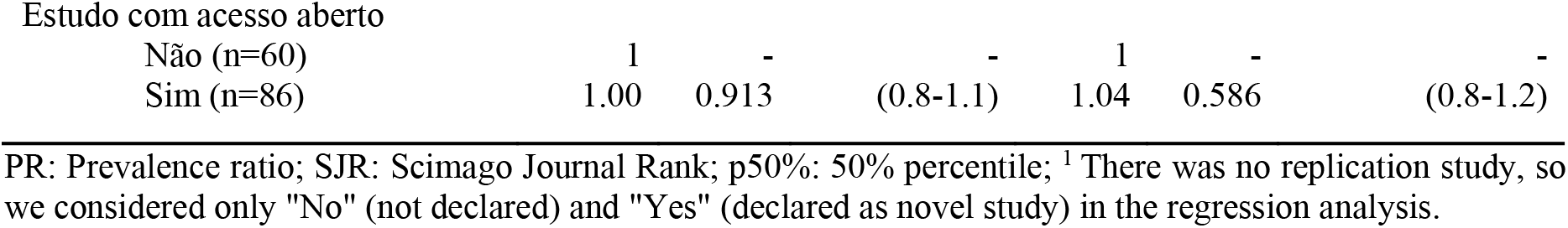
Association between general characteristics and TIDieR adherence.

| Predictors (N=146) |  |  |
| --- | --- | --- |
| Univariate Poisson Regression |  |  |
|  | PR | CI 95% |

|  |  |  |  |
| --- | --- | --- | --- |
| Intervention setting |  |  |  |
| Centre+Home (n=8) | 1 | - | - |
| Centre-based (n=106) | 1.06 | 0.71 | (0.7-1.5) |
| Home-based (n=32) | 1.13 | 0.50 | (0.7-1.6) |

Generalized Negative Binomial Regression
|  | Univariate Model |  |  | Multivariate Model |  |  |
| --- | --- | --- | --- | --- | --- | --- |
|  | PR |  | 95%CI | PR |  | 95%CI |
| RCT arms (No) |  |  |  |  |  |  |
| 1 EBI (n=54) | 1 | - | - | 1 | - | - |
| 2 EBI (n=71) | 0.84 | 0.041* | (0.7-1.0) | 0.88 | 0.135 | (0.7-1.0) |
| 3 EBI (n=21) | 0.74 | 0.025* | (0.6-0.9) | 0.71 | 0.010* | (0.5-0.9) |
| Impact factor (SJR) |  |  |  |  |  |  |
| below p50% (<1.5) | 1 | - | - | 1 | - | - |
| above p50% (>1.5) | 1.15 | 0.071 | (1.0-1.3) | 0.97 | 0.774 | (0.8-1.1) |
| Journal continent |  |  |  |  |  |  |
| Europa (n=83) | 1 | - | - | 1 | - | - |
| Americas (n=60) | 1.25 | 0.004* | (1.0-1.4) | 1.29 | 0.010* | (1.1-1.5) |
| Oceania (n=3) | 2.01 | 0.001* | (1.3-3.0) | 1.99 | 0.001* | (1.3-2.9) |
| Material availability |  |  |  |  |  |  |
| No (n=106) | 1 | - | - | 1 | - | - |
| Yes (n=40) | 1.31 | 0.001* | (1.1-1.5) | 1.26 | 0.007* | (1.1-1.5) |
| Data availability |  |  |  |  |  |  |
| No (n=90) | 1 | - | - | 1 | - | - |
| Yes (n=56) | 1.14 | 0.100 | (0.9-1.3) | 1.05 | 0.511 | (0.9-1.2) |
| Analysis scripts availability |  |  |  |  |  |  |
| No (n=143) | 1 | - | - | 1 | - | - |
| Yes (n=3) | 1.31 | 0.281 | (0.8-2.1) | 1.04 | 0.863 | (0.6-1.7) |
| Link to a protocol |  |  |  |  |  |  |
| No (n= 122) | 1 | - | - | 1 | - | - |
| Yes (n= 24) | 1.11 | 0.311 | (0.9-1.3) | 1.05 | 0.650 | (0.8-1.3) |
| (Pre)registration |  |  |  |  |  |  |
| Não (n= 32) | 1 | - | - | 1 | - | - |
| Sim (n= 114) | 0.95 | 0.611 | (0.8-1.1) | 0.91 | 0.369 | (0.7-1.1) |
| Replication or Novel study<sup>1</sup> |  |  |  |  |  |  |
| No (n= 117) | 1 | - | - | 1 | - | - |
| Yes, novel (n= 29) | 1.12 | 0.236 | (0.9-1.3) | 1.13 | 0.207 | (0.9-1.3) |
| Declara conflito de interesse |  |  |  |  |  |  |
| Não (n= 31) | 1 | - | - | 1 | - | - |
| Sim (n= 115) | 0.83 | 0.061 | (0.7 – 1.00) | 0.86 | 0.135 | (0.7-1.0) |
| Declara financiamento |  |  |  |  |  |  |
| Não (n= 22) | 1 | - | - | 1 | - | - |
| Sim (n= 124) | 0.92 | 0.468 | (0.7-1.1) | 0.88 | 0.261 | (0.7-1.1) |
| Estudo com acesso aberto |  |  |  |  |  |  |
| Não (n=60) | 1 | - | - | 1 | - | - |
| Sim (n=86) | 1.00 | 0.913 | (0.8-1.1) | 1.04 | 0.586 | (0.8-1.2) |

PR: Prevalence ratio; SJR: Scimago Journal Rank; p50%: 50% percentile; <sup>1</sup> There was no replication study, so we considered only "No" (not declared) and "Yes" (declared as novel study) in the regression analysis.

## 4. Conclusions

We found an inadequate and insufficient completeness of reporting of CVR interventions of EBI accordingly to the recommendations of the TIDieR reporting guideline. Overall, about 40% of TIDieR adherence was found, with most interventions reporting its items improperly or insufficiently. The investigation within our secondary outcomes did not shown association between the intervention care setting and the interventions’ adherence to TIDieR guideline, as well as for most of the potential predictors (in exception to continent and materials availability).

Considering TIDieR adherence, the “*Modifications*” was the less frequently reported among EBI arms analyzed. Using the criteria to “*not give the benefit of the doubt*” to authors [29], that has been adopted by the vast majority of methodological assessment and risk of bias tools (**note**: we point out we acknowledge that the TIDieR is a reporting guideline and not a methodological checklist), what was really done may have been underestimated in our study due to the poor reporting of the authors, also found in the literature [30,31]. Modifications may occur and are fair, potentially being a result of unexpected changes in study’s circumstances. Whenever happening, it is crucial that care provider expertise should be ensured to keep the study’s internal validity, and therefore tobe replicated/implemented [32]. However, only 30% of the interventions studied in our sample described it adequately, still putting interventions at a high risk to not be implemented.

In the same manner, the description of intervention schedule and intensity (*i.e*., “*When and How Much*”), is pivotal for EBIs. However, nearly half of our sample described the “*dosage”*, which Abell and colleagues reached only after the search for additional sources or contacting the corresponding author [3], although other studies showed 80% to 90% adherence of this item [25,33]. We postulate that variations and the complexity of intervention methods, settings or mode of delivery among RCTs in CVR should be acknowledged [12,18,34,35]. Also, dosis titration may interact with the natural effect of the dosis by itself – therefore, to tailor EBI is a fundamental aspect of EBI [36,37]. In our sample, only around 25% of the assessed interventions detailed tailoring procedures of the EBIs. We then found that 21% of the EBI’s arms described with proper adequation item 11 (*How well: planned*), and only 16% delivered the intervention according to the planning. These details are imperative, since EBIs are multimodal and need to cope with patients’ expectations and needs, exercise interventions outcomes vary greatly between subjects [38] and the effectiveness of an EBI is highly depent of its planning and monitoring [39–41].

Finally, we found no association between the intervention care setting and adherence to TIDieR, as well as between several other potential factors related to transparency and methodological quality of the study. Therefore, we could not state whether to deliver the intervention in clinical centers or at home or, even more, in a combined manner, are predictors of better descriptions of interventions and then, perhaps speculate the potential of third parties to mirror these interventions and provide better care to patients. Methodologically, studies with more comparator groups have less probability to adequately describe its interventions, which may be an implication of implied overload related to the reporting process – from planning to the final manuscript [42]. We should acknowledge that editorial policies may limit it whenever imposing limit for wording, which of course may impact the completeness of reporting. Authors may provide additional information through supplements or depositing materiais in a public repository.

We should disclose our perceived limitations related to this study and point out future directions. First, we did not contact authors or search for secondary publications (*e.g*., ancillary studies, registers protocols, *etc*.), which may provide further details herein pointed as absent. Second, we selected the SJR Impact Factor as a ladder to rank journals to select CVR EBI randomized clinical trials, although impact metrics does not reflect necessarily the quality of the journal and its editorial process (which reflects policies for reporting guidelines and rigour in peer-revieing) and, because of this, we may have overestimated and/or underestimated our sample in its terms. Third, the association between materials availability and TIDieR adherence may be biased by its intrinsic similarities within the collection forms or may be related to author’s intention to make the method’s materials available, which often includes intervention materials. Based in our findings, we believe that they support editorial committees to push up journals in the field for a better intervention reporting (*e.g*., ICMJE and WAME), as well as to provide empirical evidence to guide stakeholders in which levels they may intervene for this purpose. Furthermore, to reinforce those educational initiatives focusing on completeness of reporting of interventions are still needed.

We concluded that EBIs in CVR lack to report fundamental aspects of the interventions to be replicated and implemented by third parties based on their reports, using the TIDieR reporting guideline as reference. They are also not associated with intervention care settings or several methodological and transparency variables, with exceptions to the continent of publication and the materials sharing description.

## Data Availability

All data produced are available online at https://osf.io/754ht/

## 5. Acknowledgements

We thank Professor Lucas Helal, Ph.D, for his support in terms of design and methods.

## 7. Appendix

### A. Search strategy

Randomized Controlled Trial Filter [21]

(randomized controlled trial [pt] OR controlled clinical trial [pt] OR randomized controlled trials [mh] OR random allocation [mh] OR double-blind method [mh] OR single-blind method [mh] OR clinical trial [pt] OR clinical trials [mh] OR (“clinical trial” [tw]) OR ((singl* [tw] OR doubl* [tw] OR trebl* [tw] OR tripl* [tw]) AND (mask* [tw] OR blind* [tw])) OR (“latin square” [tw]) OR placebos [mh] OR placebo* [tw] OR random* [tw] OR research design [mh:noexp] OR comparative study [mh] OR evaluation studies [mh] OR follow-up studies [mh] OR prospective studies [mh] OR cross-over studies [mh] OR control* [tw] OR prospectiv* [tw] OR volunteer* [tw]) NOT (animal [mh] NOT human [mh]))

PubMed/MEDLINE

#1 (“Title of the journal” [Journal])

#2 (Cardiac Rehabilitation[Mesh] OR Exercise Therapy[Mesh] OR Sports[Mesh] OR Physical Exertion[Mesh] OR rehabilitation OR (physical* AND (fitness or training or therapy or activity)) OR Exercise[Mesh] OR (train*[tiab](strength[tiab] or aerobic[tiab] or exercise[tiab])) OR (exercise[tiab] or fitness[tiab]))

#3 (treatment[tiab] or intervention[tiab] or program[tiab])) OR Rehabilitation[Mesh] OR kinesiotherapy

#1 AND #2 AND #3

### B. Intervention description completeness assessment requirements (from TIDieR)

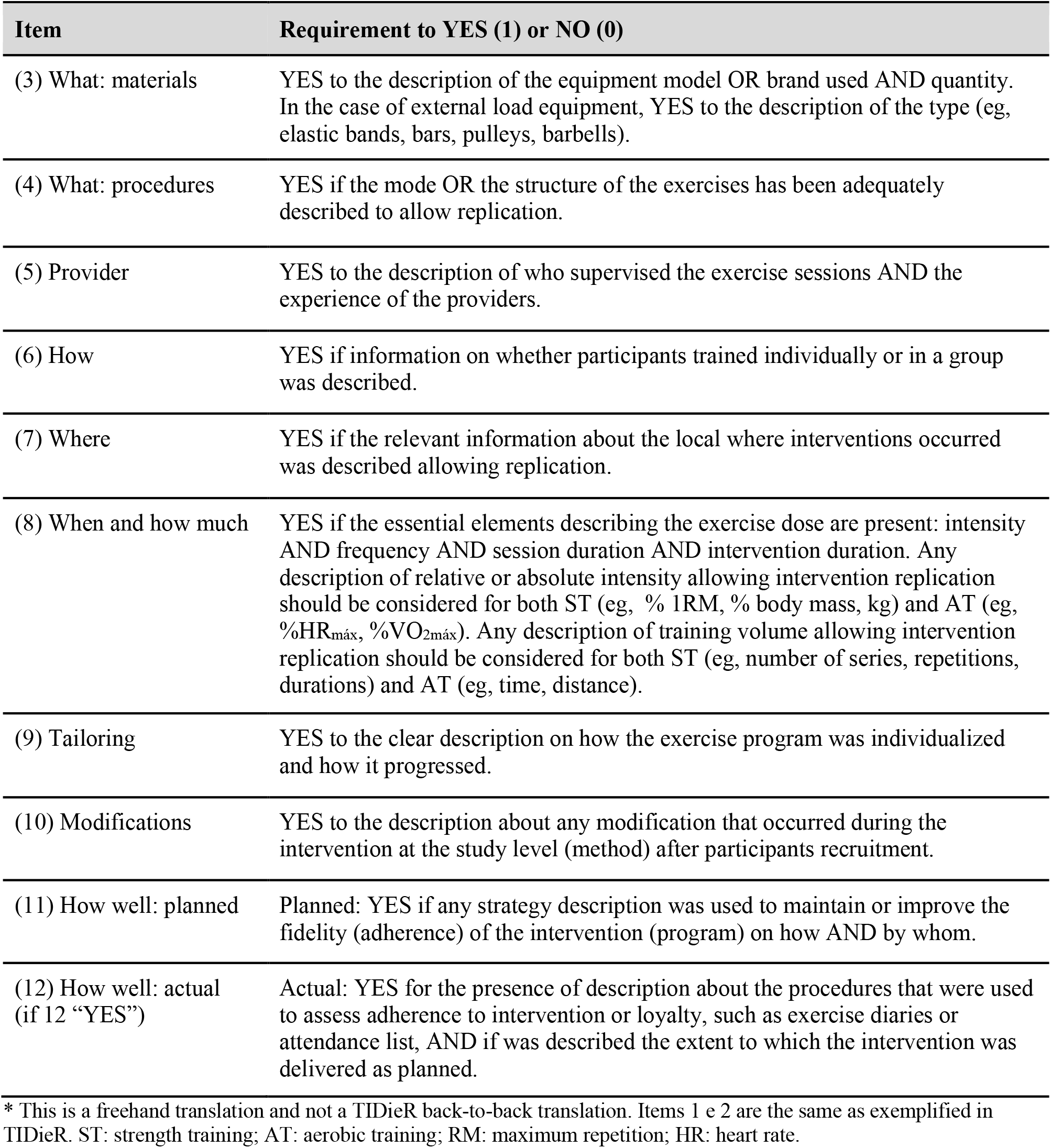

B. Intervention description completeness assessment requirements (from TIDieR).
| Item | Requirement to YES (1) or NO (0) |
| --- | --- |
| (3) What: materials | YES to the description of the equipment model OR brand used AND quantity. In the case of external load equipment, YES to the description of the type (eg, elastic bands, bars, pulleys, barbells). |
| (4) What: procedures | YES if the mode OR the structure of the exercises has been adequately described to allow replication. |
| (5) Provider | YES to the description of who supervised the exercise sessions AND the experience of the providers. |
| (6) How | YES if information on whether participants trained individually or in a group was described. |
| (7) Where | YES if the relevant information about the local where interventions occurred was described allowing replication. |
| (8) When and how much | YES if the essential elements describing the exercise dose are present: intensity AND frequency AND session duration AND intervention duration. Any description of relative or absolute intensity allowing intervention replication should be considered for both ST (eg, % 1RM, % body mass, kg) and AT (eg, %HR<sub>max</sub>, %VO<sub>2max</sub>). Any description of training volume allowing intervention replication should be considered for both ST (eg, number of series, repetitions, durations) and AT (eg, time, distance). |
| (9) Tailoring | YES to the clear description on how the exercise program was individualized and how it progressed. |
| (10) Modifications | YES to the description about any modification that occurred during the intervention at the study level (method) after participants recruitment. |
| (11) How well: planned | Planned: YES if any strategy description was used to maintain or improve the fidelity (adherence) of the intervention (program) on how AND by whom. |
| (12) How well: actual (if 12 “YES”) | Actual: YES for the presence of description about the procedures that were used to assess adherence to intervention or loyalty, such as exercise diaries or attendance list, AND if was described the extent to which the intervention was delivered as planned. |

\* This is a freehand translation and not a TIDieR back-to-back translation. Items 1 e 2 are the same as exemplified in TIDieR. ST: strength training; AT: aerobic training; RM: maximum repetition; HR: heart rate.

## Additional information

### Compliance with reproducibility and data sharing standards

This project is in accordance with the current standards of transparency indicated by the International Committee of Journal Medical Editors, the Committee of Publication Ethics, has a Principal Investigator (PI) as a signatory of the San Francisco Declaration of Research Assessment (DORA) and endorses the Hong Kong Manifesto for the assessment of researchers, faculty, and others. This project intends to publish all its scientific pieces in a pre-print version and an open-access journal, whenever possible.

Independent authors will have full access to our data including: Zotero libraries with eligible and ineligible articles, statistical codes used in analysis, statistical analysis, glossary of variables and protocol in our public repository without time constraints nor request conditions. The data will be available immediately following publication with no end date. Data is available indefinitely at our <u>OSF repository</u>.

### Data availability statement

Repository: Open Science Framework - Assessing the completeness of reporting of randomized clinical trials of exercise-based cardiovascular rehabilitation. https://osf.io/754ht/.

DOI: 10.17605/OSF.IO/754HT

This project contains the following underlying data:

- Rawdata.csv (descriptive raw data)
- Research Protocol.pdf (project’s protocol)
- Stata analysis.rtf (data analysis file)

Data are available under the terms of the **Creative Commons Zero “No rights reserved” data waiver** (CCO1.0 Public domain dedication).

### Author’s contributions

Conception: GWF

Data Curation: GWF

Formal Analysis: GWF,

Investigation: GWF, RZ, LL, ASK

Methodology: GWF

Data treatment: GWF Writing: GWF, TC, RZ

Revision: TC, RZ, ASK Final edition:

GWF Supervision: TC

Project administration: GWF

Funding acquisition: N/A.

## Footnotes

1 (Pre)registration refers to the specification of important aspects of the study (typically hypotheses, methods, and/or analysis plan) prior to commencement of the study.

